# Population-scale machine learning models estimate individual-level risk for acute respiratory infection

**DOI:** 10.64898/2026.10.23.26366351

**Authors:** Steffen Albrecht, Ewan Smith, Hui Yee Yeo, Ji Yeon Park, Asma Gul, Isabella Hills, Jo Hilder, Rachel Tester, Mamaeroa David, Elisa Weiss, Elizabeth Wilson, Maria Stubbe, Samantha Marsh, Lorraine Castelino, Nikki Turner, Anthony C Dowell, Janine Paynter

## Abstract

Summary / Abstract

**Background:** Models based on administrative population health records can help improve understanding of disease and hospitalisation risk. This understanding can support evidence-based population health policy and processes. Influenza and other acute respiratory diseases place a large, inequitable burden on the New Zealand population and health care resources. We have developed and evaluated such models using retrospective New Zealand hospitalisation records linked to other administrative data to estimate individual-level risk of hospitalisation due to acute respiratory infection.

**Methods:** We analysed linked annual records for approximately 4·85–5·36 million people in 2017–2024. Six machine learning algorithms and eight variable sets were assessed using five-fold cross-validation grouped by demographic profile. Variables comprised demographics, continuous hospital-based (M3) and pharmaceutical-based (P3) morbidity indices, and influenza vaccination status. Models trained on one year were also evaluated on data from the following year. Performance measures were the area under the receiver operating characteristic curve (AUROC) and the average precision score (APS). SHapley Additive exPlanations (SHAP) described model contributions, including paired vaccination-input changes in 2024.

**Findings:** Annual national hospitalisation prevalence ranged from 1·01% to 1·60%. With all variables, mean cross-validation AUROC and APS were 0·8430 and 0·1283 for LightGBM (LGB), and 0·8415 and 0·1285 for XGBoost (XGB). Adding both morbidity indices to the demographics increased LGB APS from 0·0720 to 0·1158; adding vaccination further increased it to 0·1283. Next-year AUROC and APS averaged 0·8429 and 0·1267 for LGB and 0·8423 and 0·1277 for XGB. Age and morbidity were the most influential variables. Among vaccinated individuals, SHAP analysis indicated that vaccination contributed most strongly towards lower model-estimated hospitalisation risk in the oldest age group.

**Interpretation:** Combining historical morbidity and vaccination information in gradient boosting models improved the ability to distinguish between people who experienced respiratory hospitalisation and those who did not in annual New Zealand population data. Similar next-year performance and interpretable variation across age and morbidity profiles support further development toward practical risk assessment, including calibration and evaluation across population groups.

**Research in context:** *Evidence before this study:* Our focused narrative review considered population-based respiratory hospitalisation models, respiratory disease burden in New Zealand, and the development of the M3 and P3 morbidity indices. Existing studies include population models for COVID-19 and infant respiratory syncytial virus, risk-factor analyses of broad respiratory admissions, and evidence that hospital and pharmaceutical morbidity measures provide complementary information.

*Added value of this study:* This is the first evaluation of national hospitalisation data linked to other national administrative data to model the risk of acute respiratory infection admission across eight annual datasets, comparing six algorithms and eight variable sets. Together with the benchmark results, these findings illustrate how gradient boosting applied to population-scale records can represent and explain combinations of routinely recorded health information.

*Implications of all the available evidence:* Quantifying historical morbidity and including vaccination information improved respiratory hospitalisation classification, with gradient boosting providing the strongest overall performance. The explanatory analyses further showed that vaccination-related contributions depended on the individual’s age and morbidity profile. Representing these clinically plausible patterns supports the use of flexible, non-linear models, such as tree-based algorithms, and rich population-scale records for developing interpretable respiratory risk assessments. The study establishes a basis for a New Zealand respiratory risk model by identifying effective algorithms and useful combinations of variables.

## Introduction

Respiratory infections are a substantial cause of hospitalisation, and their burden is unevenly distributed across the population. New Zealand studies have documented inequalities in serious infectious disease admissions and respiratory hospitalisation burden associated with age, ethnicity, and deprivation ^1,2^. Identifying people at greater risk could support prevention and guided discussions between patients and general practitioners about modifiable risk factors and mitigation strategies. Routinely collected population records allow us to investigate this vulnerability across a broad range of respiratory hospitalisation outcomes without requiring new clinical measurements and participant burden.

Many respiratory models concern outcomes after infection or presentation to healthcare ^3,4^. Population-based examples include *QCOVID* for COVID-19 hospitalisation and mortality among adults, and a registry-based model of respiratory syncytial virus (RSV) hospitalisation among infants ^5,6^. A Scottish national study also identified age, deprivation, existing conditions, and previous admissions as risk factors for a broad group of acute respiratory infection admissions ^7^. While forecasting aggregate respiratory admissions to predict healthcare demand in New Zealand has been addressed ^8^, an individual-level model to identify people at increased risk of hospitalisation is lacking. An individual model can help discern baseline risk due to unmodifiable characteristics, such as age, from modifiable factors and identify relative contributions of different factors to overall risk.

The integration of variables was informed by international literature on risk factors for severe influenza, which highlights age and underlying health conditions as important determinants of severe outcomes ^9^. Representing the combined burden of long-term conditions is therefore an important consideration in developing an individual risk model. In New Zealand, the Measuring Multimorbidity (M3) and Pharmaceutical Prescribing Profile (P3) indices summarise historical morbidity using hospital diagnoses and pharmaceutical dispensing, respectively ^10,11^. These indices offer a practical way to incorporate information about multiple conditions from routinely collected records and to assess whether the two sources provide complementary information. Influenza vaccination is also relevant as an established preventive measure against an important cause of respiratory hospitalisation and, unlike age and most medical conditions, represents a modifiable factor ^12^. Together with demographic and deprivation variables, these measures provide a starting point for establishing a population model using routinely available information. Investigating the contribution of these variables provides a foundation for subsequent models incorporating more detailed clinical, behavioural, and environmental variables.

Using New Zealand population data, we aimed to develop and evaluate a model that distinguishes between people with and without a hospital admission for respiratory infectious disease. Using linked annual population records from 2017 to 2024, we compared modelling approaches and quantified the extent to which models improved by incorporating historical morbidity and influenza vaccination data, in addition to demographic variables. We also examined whether models maintained their performance when applied to the following year and explored how age, morbidity, and vaccination contributed to model estimates.

## Methods

### Study design and data

We conducted a retrospective population-level classification study using linked records from the Stats NZ Integrated Data Infrastructure (IDI), a secure research database linking de-identified administrative and survey records about people and households in New Zealand. Annual datasets covered 2017–2024. Sources included population and address records aggregated to the mesh block level, hospital discharge information, immunisation records, and pharmaceutical dispensing records. The unit of analysis was the person within an annual dataset; individuals could appear in several years. People with missing required information were excluded before modelling, and no imputation was applied.

### Outcome and variables

The binary outcome indicated at least one qualifying hospitalisation during the calendar year, identified using a broad set of respiratory disease codes from the International Statistical Classification of Diseases and Related Health Problems, Tenth Revision, Australian Modification (ICD-10-AM). The complete code list is provided in **Table S1** (supplementary material). Codes were selected to achieve some specificity for influenza and based on previous literature ^1,13^, clinical advice and codes most commonly associated with influenza-positive SARI cases from the SHIVERS study ^14^.

Demographics comprised age, sex, and area-level deprivation score ^15^ and five non-exclusive ethnicity indicators: Asian, European, Māori, Pacific peoples, and Middle Eastern, Latin American, and African (MELAA). Individuals could identify with more than one ethnic group. All five binary ethnicity variables were retained simultaneously, giving eight demographic input columns. In tables and plots, *Europe* and *Pacific* denote European and Pacific peoples, respectively; *DEP* denotes the deprivation score, *FV* denotes the influenza vaccination status, and *Demo* denotes the demographic variable set.

To incorporate information on individual multimorbidity, we used the M3 and P3 indices, developed by Stanley and colleagues using New Zealand population health data ^10,11^. M3 and P3 indices were derived from linked IDI records and included as continuous variables. M3 used hospital diagnoses from the five years before each annual snapshot, whereas P3 used pharmaceutical dispensing records from the preceding year. Neither index included information from the outcome year. For hospitalised individuals, FV was one (*FV=1*) if vaccination occurred in the same calendar year at least 14 days before the qualifying admission. For people without hospitalisation, FV was one if vaccination occurred at any time during that year; otherwise, it was zero (*FV=0*).

Eight variable sets were evaluated: demographics alone; demographics with FV, M3, or P3 separately; demographics with M3 and P3; demographics with FV and either morbidity index; and demographics with all three. The full set contained 11 input columns.

**Table 1** summarises the characteristics of the 2024 study population overall and by hospitalisation status.

**Table 1.** Characteristics of the 2024 analysis population by respiratory hospitalisation status.

| **Characteristic** | **All individuals** | **With hospitalisation** | **Without hospitalisation** |
|---|---|---|---|
| **Number of individuals** | **5 355 519** | **85 572** | **5 269 944** |
| **Age group, years** |  |  |  |
| 0 | 51 639 (1.0 %) | 4 281 (5.0 %) | 47 355 (0.9 %) |
| 1–4 | 235 437 (4.4 %) | 13 959 (16.3 %) | 221 478 (4.2 %) |
| 5–14 | 665 415 (12.4 %) | 7 284 (8.5 %) | 658 128 (12.5 %) |
| 15–64 | 3 484 077 (65.1 %) | 26 361 (30.8 %) | 3 457 716 (65.6 %) |
| 65+ | 918 951 (17.2 %) | 33 687 (39.4 %) | 885 261 (16.8 %) |
| **Sex** |  |  |  |
| Female | 2 682 939 (50.1 %) | 42 630 (49.8 %) | 2 640 306 (50.1 %) |
| Male | 2 672 580 (49.9 %) | 42 942 (50.2 %) | 2 629 638 (49.9 %) |
| **Ethnicity** |  |  |  |
| Asian | 1 041 447 (19.4 %) | 10 245 (12.0 %) | 1 031 202 (19.6 %) |
| Europe | 3 525 792 (65.8 %) | 55 746 (65.1 %) | 3 470 046 (65.8 %) |
| Pacific | 498 327 (9.3 %) | 11 805 (13.8 %) | 486 519 (9.2 %) |
| Māori | 885 822 (16.5 %) | 19 533 (22.8 %) | 866 286 (16.4 %) |
| MELAA | 113 991 (2.1 %) | 1 425 (1.7 %) | 112 566 (2.1 %) |
| **DEP quintile** |  |  |  |
| Q1 | 1 064 223 (19.9 %) | 11 700 (13.7 %) | 1 052 523 (20.0 %) |
| Q2 | 1 042 248 (19.5 %) | 13 737 (16.1 %) | 1 028 511 (19.5 %) |
| Q3 | 1 057 434 (19.7 %) | 15 852 (18.5 %) | 1 041 585 (19.8 %) |
| Q4 | 1 088 613 (20.3 %) | 19 278 (22.5 %) | 1 069 338 (20.3 %) |
| Q5 | 1 102 998 (20.6 %) | 25 011 (29.2 %) | 1 077 990 (20.5 %) |
| **Historical morbidity** |  |  |  |
| P3 > 0 | 2 297 685 (42.9 %) | 57 081 (66.7 %) | 2 240 595 (42.5 %) |
| M3 > 0 | 642 813 (12.0 %) | 35 721 (41.7 %) | 607 086 (11.5 %) |
| **Influenza vaccination** |  |  |  |
| Recorded FV = 1 | 1 204 566 (22.5 %) | 20 655 (24.1 %) | 1 183 914 (22.5 %) |

Values are n (%) except the total number of individuals. Percentages use the total in each column. To comply with StatsNZ data confidentiality rules, counts were randomly rounded to base three; totals and percentages may not add exactly.
Ethnicity is reported using non-exclusive indicators; individuals may appear in more than one group, so ethnicity percentages may sum to more than 100%. Europe and Pacific denote European and Pacific peoples, respectively. MELAA denotes Middle Eastern, Latin American, and African; DEP denotes deprivation score; FV denotes influenza vaccination status. M3 and P3 are the hospital-based and pharmaceutical-based morbidity indices, respectively.
For people with hospitalisation, FV = 1 denotes vaccination within the same year at least 14 days before the qualifying admission; for others, it denotes vaccination at any time during that year.

### Algorithms and cross-validation

We treated hospitalisation as a binary classification task: each annual record was labelled as having at least one qualifying hospital admission (1) or not (0). During training, machine learning classification algorithms learned relationships between the recorded variables and these labels. Each fitted model then produced a continuous score between 0 and 1 for the hospitalisation class, with higher scores indicating greater model-estimated hospitalisation risk. We evaluated how well these scores distinguished people with and without hospitalisation, without selecting a threshold to assign individuals to either class. We used scores from the class-balanced models to rank individuals by hospitalisation risk and did not assess calibration.

We compared logistic regression (LR), random forest (RF), LightGBM (LGB), XGBoost (XGB), and single-tree versions of LightGBM and XGBoost, labelled Tree (lgb) and Tree (xgb) ^16–18^. RF combines the outputs of multiple decision trees, whereas LGB and XGB build trees sequentially, with each additional tree refining the combined model. The ensembles used 100 trees (the default parameter), and the single-tree versions used 1 boosting iteration. LR included linear terms for continuous variables on the log-odds scale and no interaction terms, serving as a linear baseline model. Decision-tree models can accommodate nonlinear relationships and interactions without specifying these patterns in advance. Analyses were conducted on the raw individual-level data in secure Stats NZ Data Labs. Computational constraints in the Data Lab informed the choice of algorithms and fixed settings; no hyperparameter tuning was performed (see supplementary material **Table S2**).

Five non-overlapping test folds were formed within each year using demographic-profile groups. A profile combined age band, sex, the five ethnicity indicators, and a 10-step deprivation band. One representative was randomly selected from each profile, and a five-fold stratified allocation was applied to these representatives based on their outcomes. All individuals with a given profile were assigned to the same fold. Each fold was held out once, with approximately 80% of annual records used for training and 20% for testing. This profile grouping was applied to ensure that multiple samples with the same or very similar combinations of variable values are represented in both the training and testing sets, which can be an issue for the model when using only demographic variables, as these are mainly categorical. The same folds were used across algorithms and variable sets within the cross-validation.

Variables were standardised using means and standard deviations fitted to each training set, then transformed in the held-out data. Class balancing was applied during training, using the appropriate algorithm parameters. LR used balanced class weights; XGB used the negative-to-positive training count ratio; LGB used its class-imbalance option; and RF used corresponding observation weights.

### Performance and temporal evaluation

Hospitalisation caused by respiratory infection, whilst making a large contribution to seasonal hospital admissions, is infrequent at the population level. In the annual datasets, approximately 1·5% to 1·6% of people had a qualifying admission in most years (**Table 2**). Most people do not have a hospital admission. This is termed a *class imbalance*, which makes overall classification accuracy uninformative on its own. For example, a naïve model that always predicts no hospitalisation achieves an accuracy of 98·5% at a prevalence of 1·5%. Therefore, we use threshold-independent performance measures to evaluate model performance and focus on the average precision score (APS), as the area under the receiver operating characteristic curve (AUROC) can be misleading when class imbalance is high ^19^.

**Table 2.** Annual analysis of populations and hospitalisation prevalence.

| Year | People | Hospitalised people | Prevalence |
|---|---|---|---|
| 2017 | 4 851 537 | 77 319 | 1·59% |
| 2018 | 4 933 908 | 75 825 | 1·54% |
| 2019 | 5 011 356 | 78 888 | 1·57% |
| 2020 | 5 051 160 | 51 057 | 1·01% |
| 2021 | 5 090 370 | 60 819 | 1·19% |
| 2022 | 5 141 100 | 77 556 | 1·51% |
| 2023 | 5 264 985 | 83 181 | 1·58% |
| 2024 | 5 355 519 | 85 572 | 1·60% |

Counts are based on released cross-validation population totals and were randomly rounded to base three. Annual populations overlap and should not be summed as unique individuals.

Fold scores were averaged within each year, and annual means were weighted equally. These summaries average evaluation scores rather than pooling individual model outputs. The algorithms were first compared using the full variable set; variable-set comparisons then focused on LGB and XGB. Increments from adding FV were matched by year and fold against the corresponding set without FV.

For next-year evaluation, models trained on complete annual datasets from 2017 through 2023 were applied to the following year’s population. Training-year transformations and class weights were retained. Scores were averaged equally across seven year-pairs and compared with cross-validation results for the same evaluation years. This assesses temporal transfer within the national data system, where individuals can overlap between years. Overall performance summaries included all eight cross-validation years (2017–2024) and all seven next-year evaluations (supplementary **Tables S3–S6**). **Figure 2** compares variable sets after excluding the evaluation years 2020 and 2021, which were influenced by pandemic-related changes in respiratory virus circulation and markedly lower hospitalisation prevalence.

### Variable importance and SHAP

For LGB and XGB, gain importance represented the total split gain attributable to each variable, and split importance represented its frequency of use ^17,18^. Values were normalised separately within each fitted model and importance type, then averaged across folds and years and expressed as percentages.

SHapley Additive exPlanations (SHAP) describe how each variable contributes to an individual model-estimated hospitalisation risk. All variable contributions, together with a model-specific reference value, sum to the model estimate (on the log-odds scale) ^20^. For this analysis, the original models and real records inside the Data Lab were used. SHAP values were derived for a subset of 1,000,000 people randomly selected from the full dataset for the same year or the next year. A subset was used to reduce computational resource requirements. Based on the small variation observed in model performance across cross-validation folds, we expect that a random subset covering ∼20% of the data reflects the distributions of the full dataset.

To visualise the SHAP analyses of the models, we used *beeswarm* plots (**Figure 5** and Supplementary **Figure S1**). These allowed us to examine the magnitude and direction of variable contributions on the log-odds scale.

Note that we selected LGB for the SHAP analysis, as this is the model for which we, together with Stats NZ, found a reliable workflow to release trained models after confidentiality processing. Besides XGB, LGB achieved the highest model performance scores. We used *TreeExplainer*, an efficient implementation for deriving SHAP values from tree-based models ^20^.

### Paired vaccination SHAP analysis

Among people with observed FV=1 in 2024, we conducted a descriptive paired analysis using the full-variable LGB model fitted to the complete 2024 analysis dataset. Each individual was evaluated twice with *TreeExplainer*, with FV set to one and zero, holding all other variables, the fitted model, and explainer settings fixed. Differences were defined as SHAP with FV=1 minus SHAP with FV=0. Negative values indicate lower contributions under FV=1. We summarised FV and age differences by medians and quartiles within five age groups, labelled 0–1, 1– 5, 5–15, 15–65, and 65+, overall and separately for M3=P3=0 and M3 ≠ 0 or P3 ≠ 0. Both indices remained continuous model variables. This analysis describes the fitted model’s response in the training records and does not estimate causal effects of vaccination.

## Results

### Population and algorithm performance

Annual analysis populations ranged from 4,851,537 in 2017 to 5,355,516 in 2024 (**Table 2**). Between 51,057 and 85,575 people had a qualifying hospitalisation each year. Prevalence was 1·51–1·60% in 2017–2019 and 2022– 2024, compared with 1·01% in 2020 and 1·19% in 2021.

Characteristics of the 2024 analysis population are presented overall and by respiratory hospitalisation status in **Table 1**. Compared with individuals without qualifying hospitalisation, hospitalised individuals were more frequently aged 65 years or older (39·4 % versus 16·8 %) and belonged to the highest deprivation quintile (29·2 % versus 20·5 %). A greater proportion also had M3 > 0 (41·7 % versus 11·5 %) or P3 > 0 (66·7 % versus 42·5 %).

LGB and XGB had the strongest average performance with the full set of variables (**Figure 1**; supplementary **Table S3**). LGB achieved mean AUROC 0·8430 and APS 0·1283, compared with 0·8415 and 0·1285 for XGB. Tree (xgb) achieved AUROC 0·8244 and APS 0·1054, and Tree (lgb) achieved 0·8272 and 0·0811. RF had higher AUROC than LR (0·8240 vs 0·7589), but lower APS (0·0720 vs 0·0748).

**Figure 1.**
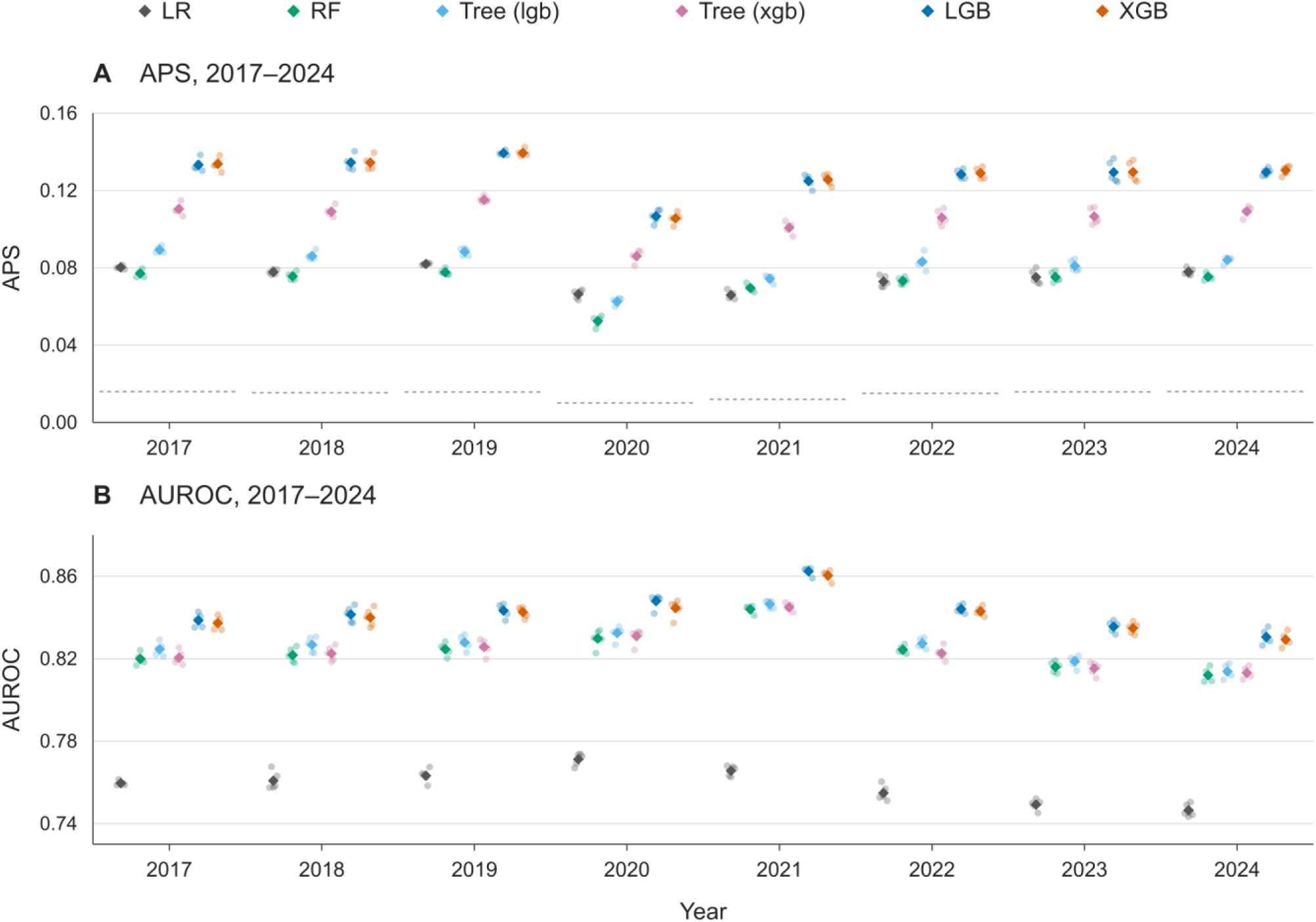
Algorithm performance across years. Average precision score (APS; A) and area under the receiver operating characteristic curve (AUROC; B) for six algorithms using the full variable set (Demo, FV, M3, and P3), covering 2017–2024. Small points show the five held-out fold scores. Diamonds indicate the mean across the five folds. Dotted segments in the APS panel indicate the approximate annual prevalence of outcomes as a baseline for the APS. Algorithm colours are consistent across panels. LGB, LightGBM; XGB, XGBoost; LR, logistic regression; RF, random forest; Tree (lgb) and Tree (xgb), single-tree LightGBM and XGBoost, respectively. Demo comprises age, sex, deprivation and five non-exclusive ethnicity indicators; FV denotes influenza vaccination status. M3 and P3 are continuous historical morbidity indices.

The APS achieved by LGB was highest in 2019 (0·1394) and lowest in 2020 (0·1067), whereas its highest AUROC occurred in 2021 (0·8624). XGB showed the same pattern, with corresponding values of 0·1395, 0·1056, and 0·8603. Comparing 2017–2019 with 2022–2024, mean LGB APS was 0·1357 versus 0·1291, and AUROC was 0·8411 versus 0·8367. XGB showed similar period differences. Mean hospitalisation prevalence was almost unchanged (1·568% vs 1·562%).

### Contribution of morbidity and vaccination

Both morbidity indices improved performance across all years (**Figure 2** and supplementary material **Tables S4 and S5**). For LGB, demographics alone yielded an AUROC of 0·7856 and an APS of 0·0720. Adding M3 increased these to 0·8259 and 0·1034; adding P3 increased them to 0·8297 and 0·0998. Including both indices yielded an AUROC of 0·8400 and an APS of 0·1158. XGB followed the same pattern: AUROC increased from 0·7830 to 0·8382 and APS from 0·0713 to 0·1154 with both indices. Relative APS increases were 60·8% for LGB and 61·9% for XGB.

**Figure 2.**
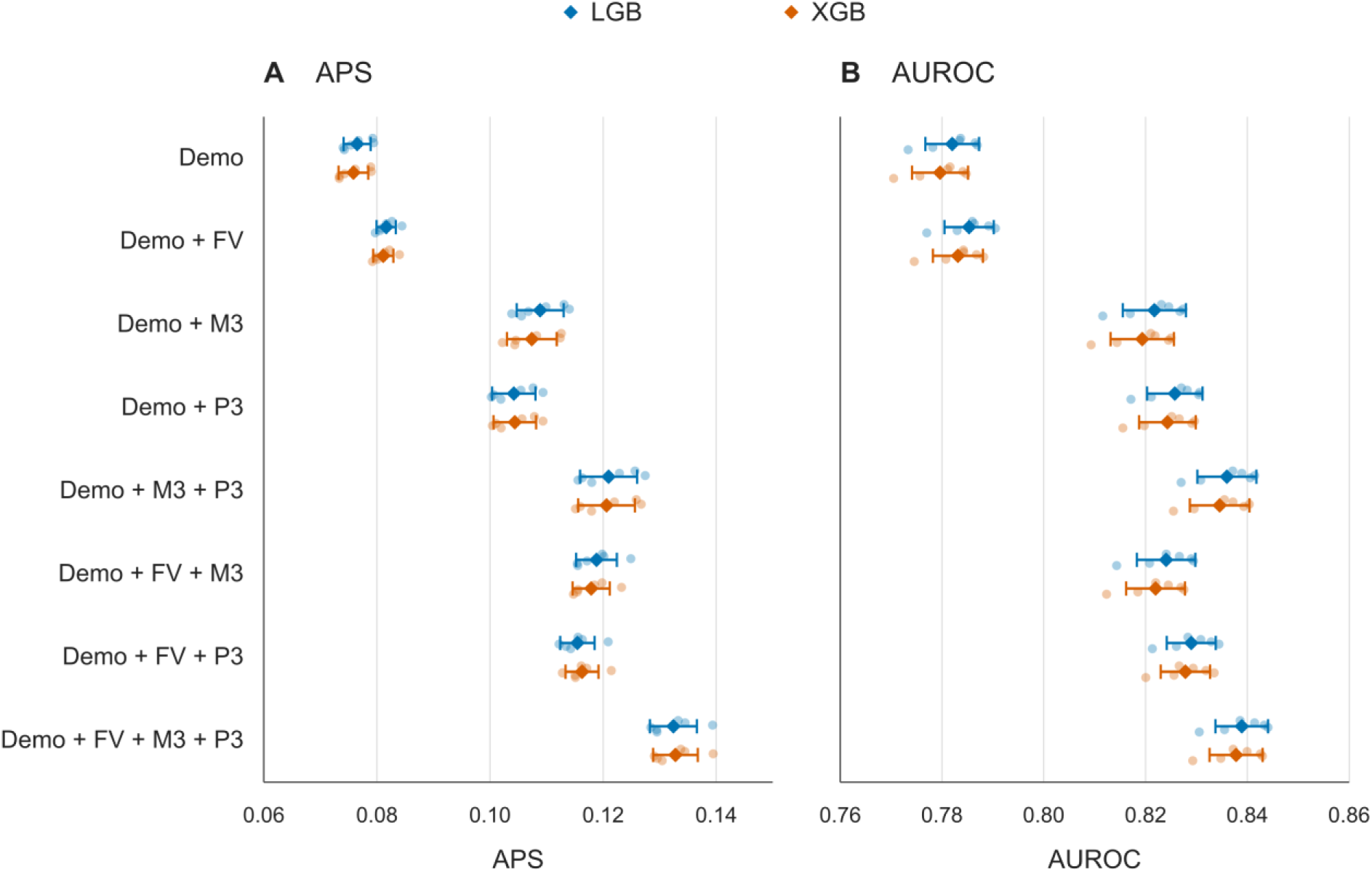
Added value of variable information. Cross-validation APS (A) and AUROC (B) for eight variable sets in LGB (blue) and XGB (orange). This figure excludes 2020 and 2021. Small points represent the six annual means from 2017–2019 and 2022–2024, each calculated from five held-out folds. Diamonds indicate the equal-year mean across those six years, and horizontal bars indicate one sample standard deviation across their annual means, not confidence intervals. LGB and XGB are vertically offset within each variable row for readability. Demo comprises age, sex, deprivation and five non-exclusive ethnicity indicators. FV denotes influenza vaccination status; M3 and P3 are continuous historical morbidity indices. LGB, LightGBM; XGB, XGBoost; APS, average precision score; AUROC, area under the receiver operating characteristic curve.

The added value of FV depended on the other variables. For LGB, FV increased APS by 0·0054 with demographics alone and by 0·0124 when both indices were included. XGB increments were 0·0055 and 0·0131, respectively. Gains with either index separately were intermediate (**Table S6**, see supplementary material).

### Gain and split importance

Age, M3, and P3 accounted for 96·2% of LGB gain and 90·4% of XGB gain with all variables (**Figure 3**; supplementary **Table S7**). Their mean LGB gain shares were 44·69%, 36·58%, and 14·91%, respectively. Age contributed the largest share in seven years; M3 ranked first in 2020. P3 ranked third in every annual LGB gain summary. Deprivation accounted for 15·98% of LGB splits and 24·15% of XGB splits, compared with gain shares of 1·45% and 3·74%. FV accounted for 1·12% of LGB gain and 3·14% of splits.

**Figure 3.**
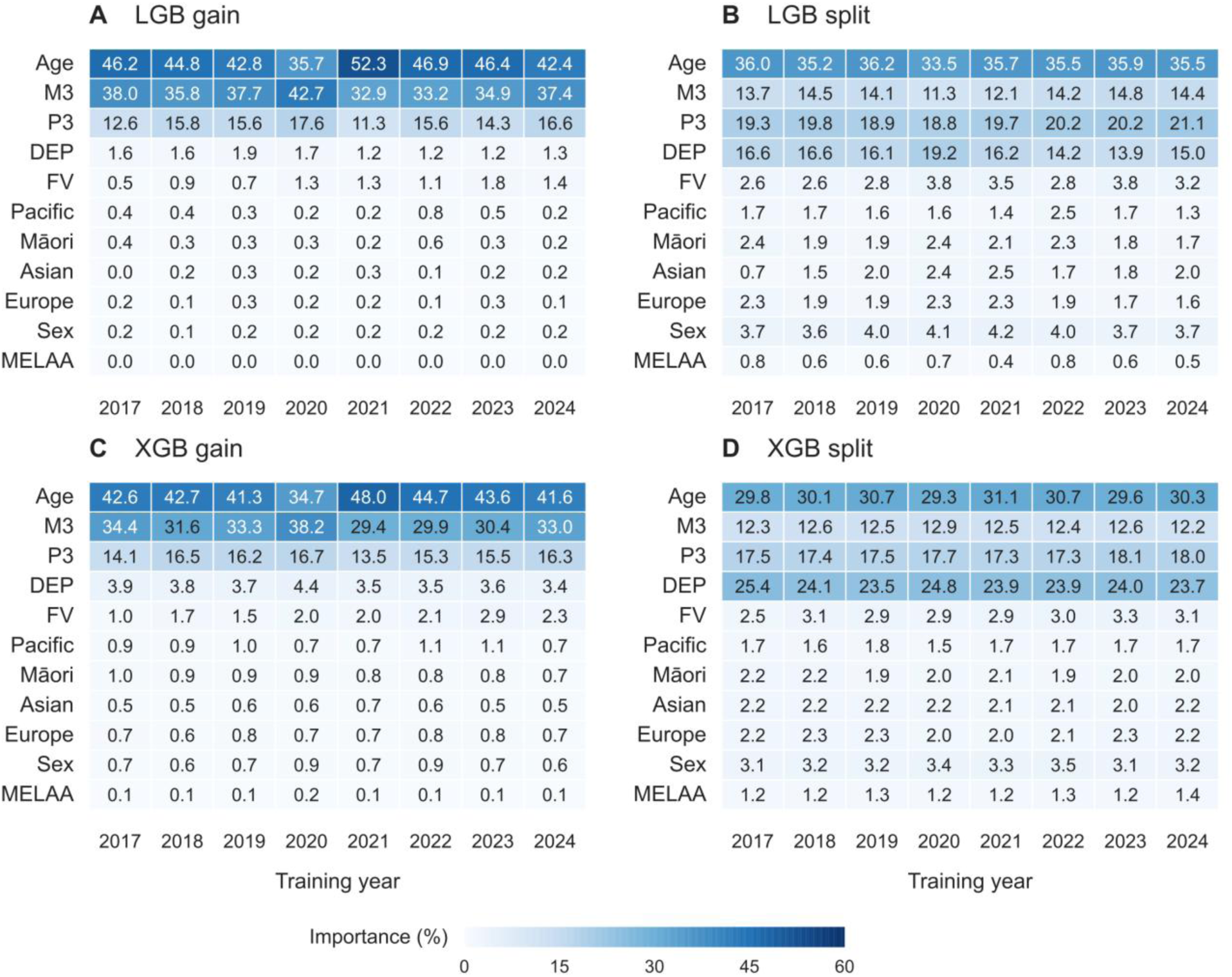
Gain and split importance across years. Normalised gain and split importance in full-variable LGB (A–B) and XGB (C–D) models by training year. Each cell shows the mean across five cross-validation fits, expressed as a percentage. Gain denotes total gain accumulated over splits involving the variable; split importance denotes split frequency. Each importance type is normalised separately within each fitted model, so each algorithm-year column sums to 100% before rounding. All panels use the same 0–60% colour scale. Variable order is held constant to facilitate comparison. Importance describes allocation within the fitted model, not a causal effect or subgroup performance. LGB, LightGBM; XGB, XGBoost; DEP, deprivation score; FV, influenza vaccination status. Europe and Pacific are shortened plot labels for the European and Pacific peoples ethnic groups, respectively. MELAA denotes Middle Eastern, Latin American, and African. Ethnic group indicators are non-exclusive.

### Next year performance and SHAP profiles

Next-year AUROC and APS averaged 0·8429 and 0·1267 for LGB and 0·8423 and 0·1277 for XGB. Aligned to the same evaluation years, mean next-year minus cross-validation differences were −0·0007 for AUROC and −0·0009 for APS for LGB, and +0·0003 and −0·0001 for XGB (**Figure 4**). Improvements from morbidity and FV remained apparent in next-year evaluations (supplementary **Tables S3–S6**).

**Figure 4.**
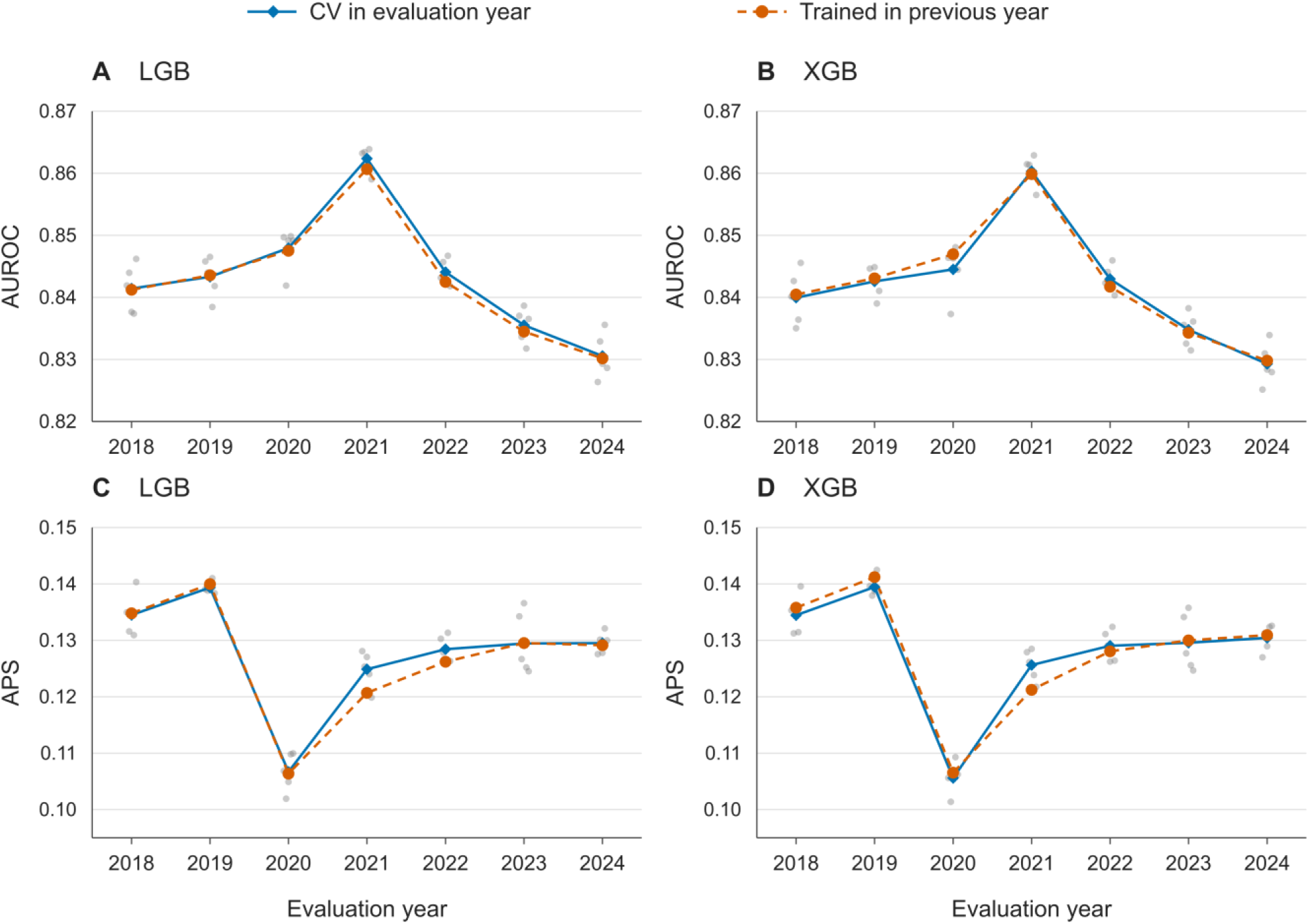
Cross validation and next year evaluation. AUROC (A–B) and APS (C–D) for full-variable LGB and XGB models, aligned by evaluation year. Blue diamonds show the mean of five cross-validation folds from that evaluation year; small grey points show individual fold scores. Orange circles show performance of a model trained on the complete preceding-year dataset and applied to the evaluation year. Each orange point is one temporal evaluation, not a mean over folds. Panels use common metric-specific limits across algorithms. Connecting lines describe annual variation. LGB, LightGBM; XGB, XGBoost; APS, average precision score; AUROC, area under the receiver operating characteristic curve.

Across the beeswarm plots, age, P3, and M3 occupied the first three displayed positions. Both young and older ages were among the positive contributions, as expected. Higher morbidity indices and deprivation generally contributed positively, whereas recorded vaccination generally contributed negatively. **Figure 5** compares next-year and same-year explanations for 2018 and 2024 (see supplementary material, **Figure S1**, for profiles derived from other years). Broad patterns were similar, although the ordering and contributions of some variables varied.

**Figure 5.**
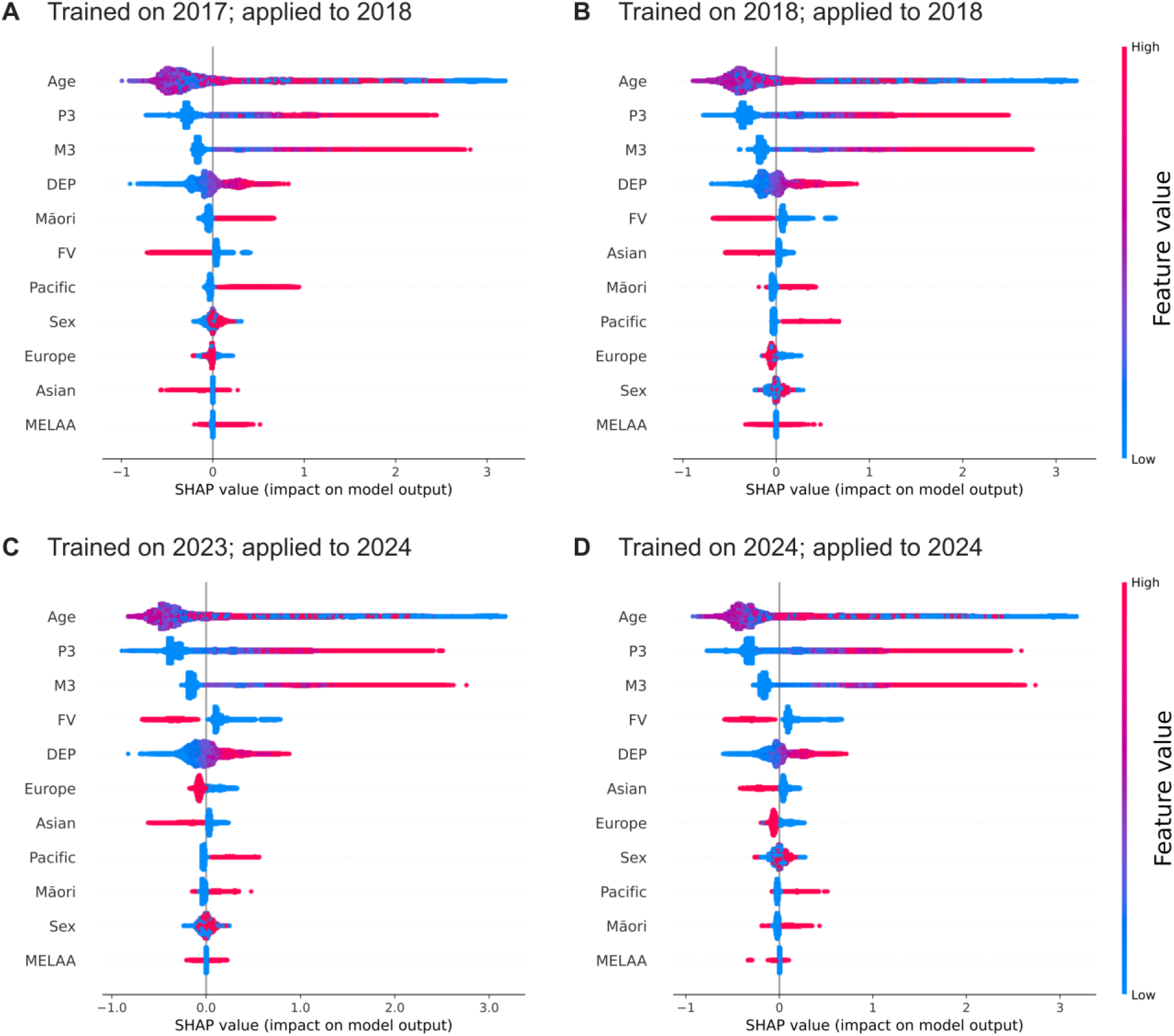
SHAP profiles in the same year and the next year application. SHapley Additive exPlanations (SHAP) beeswarm plots for full-variable models trained in 2017 and applied to 2018 (A), trained and applied in 2018 (B), trained in 2023 and applied to 2024 (C), and trained and applied in 2024 (D). Each panel represents a random sample of 1,000,000 people. Each dot shows one person’s SHAP contribution for the variable. Variables are ranked within each panel by their mean absolute SHAP value, placing those with the largest average contribution magnitudes at the top. Horizontal position indicates the contribution on the log-odds scale: positive values contribute towards higher model-estimated log-odds of hospitalisation, and negative values towards lower log-odds, relative to the model-specific reference value. Colour indicates the scaled variable’s value, from low (blue) to high (red). Values are scaled to enable a shared colour bar. For FV and ethnicity indicators, blue represents 0 and red represents 1. Greater vertical spread indicates more people with similar SHAP values. Each panel retains its own horizontal axis scale. Explanations used the original fitted models and real records inside the Data Lab. Same-year panels describe models fitted to the same annual dataset and are not held-out validation results as proposed by Lundberg *et al.* SHAP values describe model contributions, not causal effects or percentage-point changes in hospitalisation probability. Additional annual applications are presented in the supplementary material (**Figure S1)**. FV, influenza vaccination status; DEP, deprivation score; M3, hospital-based morbidity index; P3, pharmaceutical-based morbidity index. Europe and Pacific are shortened labels for European and Pacific peoples, respectively. MELAA denotes Middle Eastern, Latin American, and African. Ethnicity indicators are non-exclusive.

### Paired vaccination SHAP analysis

Setting FV to one produced a negative median change in the FV contribution across all displayed age groups (**Figure 6**). Among all vaccinated individuals within each group, median changes ranged from −0·327 to −0·418 in the four younger groups and reached −0·656 in the 65+ group. Median age-contribution changes were close to zero in younger groups (−0·001 to 0·029), compared with −0·171 in the 65+ group. Within that oldest group, changes were more negative when either morbidity index was nonzero than when both were zero: median FV changes were −0·675 versus −0·527, and age changes were −0·177 versus −0·145. All differences are in model-output units, not in model-estimated probability.

**Figure 6.**
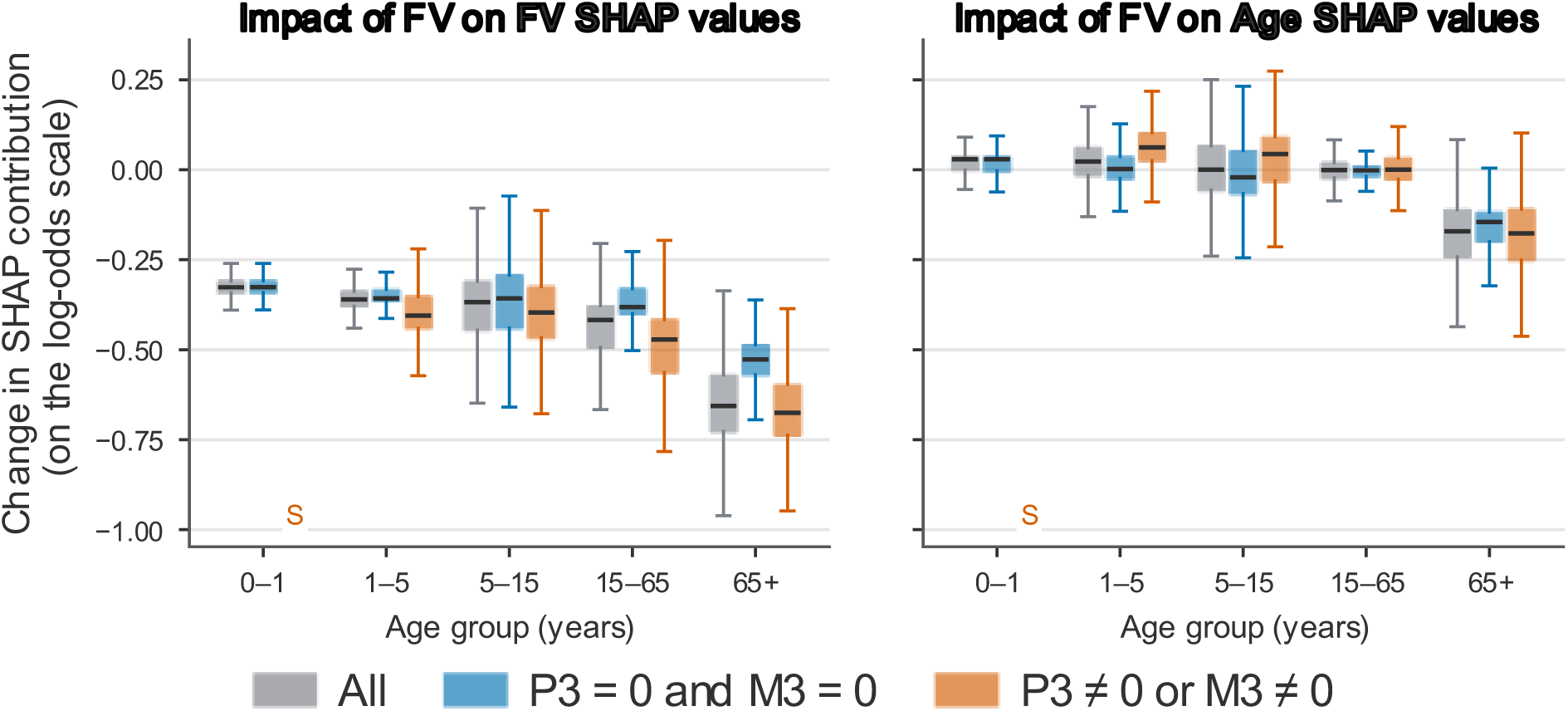
Changes in FV and age SHAP contributions under vaccination input changes. Boxplots show within-person changes in the SHAP contributions of influenza vaccination status (FV, left) and age (right) using the full-variable LGB model fitted to the complete 2024 dataset. Each individual with recorded vaccination was evaluated twice with the same model and explainer, setting FV to 1 and then to 0 while holding all other variables fixed. Differences were calculated as SHAP(FV=1) minus SHAP(FV=0) and are expressed in log-odds units in both panels. Negative values indicate a lower contribution when FV was set to 1. Each box spans the first to third quartiles (Q1 to Q3), with a line at the median. Whiskers extend to Q1 − 1·5 × IQR and Q3 + 1·5 × IQR, where IQR = Q3 − Q1. Within each age group, grey shows all vaccinated individuals, including both morbidity strata. Blue shows those with both M3 and P3 equal to zero, and orange shows those with either index nonzero. M3 and P3 were continuous model variables derived from historical hospital diagnoses and pharmaceutical dispensing, respectively. S marks a summary suppressed from Stats NZ data output due to a small subgroup size. These paired changes describe the fitted model’s response in the training records and do not estimate causal effects of vaccination.

## Discussion

Using linked New Zealand population records, we compared classification algorithms and quantified the extent to which historical morbidity and influenza vaccination information improved the ability to distinguish people with and without respiratory hospitalisation. Integrating hospital- and pharmaceutical-based morbidity indices improves models. Furthermore, our results show that these indices are complementary, with the highest APS achieved when models incorporate both. Additionally, recorded influenza vaccination provided a further improvement, with the greatest effect when both morbidity indices were included.

With all variables included, Tree (xgb) achieved a higher mean APS than RF (0·1054 vs 0·0720) despite a similar mean AUROC (0·8244 vs 0·8240). While the YDF random forest always uses a random subset of variables at each split, potentially omitting the important ones, the single XGB tree could consider all variables at every split. This difference may partly explain the single tree’s higher APS, while successive trees in the XGB and LGB ensembles could further refine the ranking of people at elevated risk.

The paired vaccination analysis showed how gradient boosting captured clinically plausible joint patterns in our data. For each vaccinated person in the 2024 analysis, we changed only the vaccination input. Recording vaccination as present produced the largest median downward shifts in both the vaccination and age contributions among people aged 65 years and older, particularly those with a nonzero morbidity index (at least one medical condition was present). Even though a person’s age is not changed in this analysis, the model assigns different contributions to the age variable based on vaccination status. This age-morbidity pattern is consistent with the priority given to older adults and people with chronic conditions in influenza vaccination recommendations. ^12^

Studies of vaccine effectiveness also document variation by age and, among older adults, by condition. ^21,22^ The SHAP changes quantify how the fitted model used vaccination information. However, estimating vaccine effectiveness within these groups requires a causal analysis.

Age and morbidity helped identify groups vulnerable to severe respiratory illness, but exposure to infection also matters. Many people with increased estimated risk had no admissions in any of the datasets. As people with admissions made up only a small fraction of the population, even a low false-positive rate could produce many false positives relative to true positives. This reduces precision and helps explain why a model can achieve good AUROC alongside modest APS. ^19^ Accordingly, in 2020, APS fell as the prevalence of qualifying admissions dropped to 1·01%. Lockdowns, border restrictions and physical distancing curtailed transmission, ^23^ and surveillance documented changes in the circulation of influenza and RSV during 2020 and 2021. ^24,25^ People vulnerable because of age or chronic illness may have had fewer opportunities to become infected. Lower prevalence itself also reduces precision at a given sensitivity and false-positive rate. The proportion with a qualifying admission rose from 1·19% in 2021 to 1·51% in 2022. During New Zealand’s Omicron waves in 2022, older adults had higher COVID-19 hospitalisation rates. ^26^ Our study may have included some of these admissions through a qualifying respiratory diagnosis such as viral pneumonia. This suggests some overlap in vulnerability across infections, although the combined outcome prevents us from attributing the rebound to a particular virus.

Compared to the pre-pandemic era, the mean APS achieved by gradient boosting was slightly lower in 2022– 2024, despite similar hospitalisation prevalence. Auckland surveillance documented changes in influenza admission rates and the age profile of admitted adults after restrictions eased, with a different pattern for human metapneumovirus. ^2^ Changes in circulating pathogens and affected populations may therefore have influenced classification, even when models were refitted each year. Similar average performance in next-year evaluations supports annual model updates, accompanied by regular assessment as respiratory disease patterns change.

Further development could use linked survey and census records to assess whether information on smoking, household crowding and employment improves classification. This is worth testing because APS remained well below its maximum of 1·0, which indicates that many variables are missing that could help a model to capture the high-risk group more precisely.

To make the models easier to use during consultations, we could replace M3 and P3 with a short list of conditions that clinicians can readily identify. Evaluating models based on individual medical conditions rather than the morbidity indices would show whether this can be done without decreasing performance. However, before the model can support individual health advice, calibration should establish how closely its scores agree with observed hospitalisation rates, with recalibration where needed. This is particularly relevant given the low prevalence rates. ^27^ Calibration, sensitivity and positive predictive value at clinically relevant thresholds should be assessed across age, ethnicity and deprivation groups, including Māori and Pacific peoples. Comparisons with models trained within these groups could show whether more specialised models add value.

Clinicians and patients could then assess whether individual explanations support informed discussions about respiratory risk and prevention.

## Data Availability

The individual-level data on which we trained all models are held in the Stats NZ Integrated Data Infrastructure and cannot be shared publicly. Access is subject to Stats NZ approval and its secure-access conditions.
Other, aggregated statistics have been released from the Stats NZ Data Labs with the following disclaimer:
Access to the data used in this study was provided by Stats NZ under conditions designed to give effect to the security and confidentiality provisions of the Data and Statistics Act 2022. The results presented in this study are the work of the authors, not Stats NZ or individual data suppliers. These results are not official statistics. They have been created for research purposes from the Integrated Data Infrastructure (IDI), which is carefully managed by Stats NZ.

## Author contributions

**SA:** conceptualisation, methodology, data curation, validation, formal analysis, investigation, writing manuscript, supervision. **ES:** conceptualisation, data acquisition, methodology, validation, formal analysis, investigation, writing manuscript. **HYY:** conceptualisation, validation. **JYP:** conceptualisation, validation. **AG:** conceptualisation, validation. **IH:** conceptualisation, validation. **JH:** conceptualisation, validation. **RT:** conceptualisation, validation. **MD:** conceptualisation, validation. **E Weiss:** conceptualisation, validation. **E Wilson:** conceptualisation, validation. **MS:** conceptualisation, validation. **SM:** conceptualisation, validation. **LC:** conceptualisation, validation, supervision. **NT:** conceptualisation, validation, supervision. **ACD:** conceptualisation, validation, supervision. **JP:** conceptualisation, data acquisition, methodology, validation, formal analysis, investigation, writing manuscript, supervision. **All authors** reviewed and edited the manuscript.

## Acknowledgments

Our grateful thanks to Flu Lab for funding the Triple R team’s research in Risk, Resilience, and Response (RRR): Mitigating the Impact of Influenza and other Respiratory Viruses in Aotearoa/New Zealand, of which this project is a part.

## Ethic declarations

The usage of all data involved in this study has been approved by the AUCKLAND HEALTH RESEARCH ETHICS COMMITTEE (AHREC), University of Auckland, New Zealand, Approval Ref. AH27716.

Access to the data used in this study was provided by Stats NZ under conditions designed to give effect to the security and confidentiality provisions of the Data and Statistics Act 2022. The results presented in this study are the work of the authors, not Stats NZ or individual data suppliers. These results are not official statistics. They have been created for research purposes from the Integrated Data Infrastructure (IDI), which is carefully managed by Stats NZ.

## Data sharing

The individual-level data on which we trained all models are held in the Stats NZ Integrated Data Infrastructure and cannot be shared publicly. Access is subject to Stats NZ approval and its secure-access conditions.

## Declaration of generative AI use

OpenAI Codex (GPT-6.1 Sol and GPT-6 Astra) was used to assist with manuscript editing, summarising released aggregate results, and preparing figures.

