## supplementary for "Population-scale machine learning models estimate individual-level risk for acute respiratory infection"

### Supplementary material for respiratory hospitalisation classification

#### Contents

**Table S1** – ICD-10-AM codes used to identify respiratory infectious disease hospitalisations

**Table S2** – Model implementations and settings

**Table S3** – Full variable model performance

**Table S4** – Average precision by variable set

**Table S5** – AUROC by variable set

**Table S6** – Increments associated with vaccination

**Table S7** – Mean gain and split importance

**Figure S1** – SHAP profiles for the additional application years

**Table S1 – ICD-10-AM codes used to identify respiratory infectious disease hospitalisations**

| Code | Description (ICD-10-AM) |
|---|---|
| B349 | Viral infection, unspecified |
| B974 | Respiratory syncytial virus as the cause of diseases classified to other chapters |
| B978 | Other viral agents as the cause of diseases classified to other chapters |
| J00 | Acute nasopharyngitis [common cold] |
| J010 | Acute maxillary sinusitis |
| J011 | Acute frontal sinusitis |
| J012 | Acute ethmoidal sinusitis |
| J013 | Acute sphenoidal sinusitis |
| J014 | Acute pansinusitis |
| J018 | Other acute sinusitis |
| J019 | Acute sinusitis, unspecified |
| J020 | Streptococcal pharyngitis |
| J028 | Acute pharyngitis due to other specified organisms |
| J029 | Acute pharyngitis, unspecified |
| J030 | Streptococcal tonsillitis |
| J038 | Acute tonsillitis due to other specified organisms |
| J039 | Acute tonsillitis, unspecified |
| J040 | Acute laryngitis |
| J041 | Acute tracheitis |
| J042 | Acute laryngotracheitis |
| J050 | Acute obstructive laryngitis [croup] |
| J051 | Acute epiglottitis |
| J060 | Acute laryngopharyngitis |
| J068 | Other acute upper respiratory infections of multiple sites |
| J069 | Acute upper respiratory infection, unspecified |
| J09 | Influenza due to identified zoonotic or pandemic influenza virus |
| J10 | Influenza due to other identified influenza virus |
| J100 | Influenza with pneumonia, other influenza virus identified |
| J101 | Influenza with other respiratory manifestations, other influenza virus identified |
| J108 | Influenza with other manifestations, other influenza virus identified |
| J11 | Influenza, virus not identified |
| J110 | Influenza with pneumonia, virus not identified |
| J111 | Influenza with other respiratory manifestations, virus not identified |
| J118 | Influenza with other manifestations, virus not identified |
| J12 | Viral pneumonia, not elsewhere classified |
| J120 | Adenoviral pneumonia |
| J121 | Respiratory syncytial virus pneumonia |
| J122 | Parainfluenza virus pneumonia |
| J123 | Human metapneumovirus pneumonia |
| J128 | Other viral pneumonia |
| J129 | Viral pneumonia, unspecified |
| J13 | Pneumonia due to Streptococcus pneumoniae |
| J14 | Pneumonia due to Haemophilus influenzae |
| J150 | Pneumonia due to Klebsiella pneumoniae |
| J151 | Pneumonia due to Pseudomonas |
| J152 | Pneumonia due to staphylococcus |
| J153 | Pneumonia due to streptococcus, group B |
| J154 | Pneumonia due to other streptococci |
| J155 | Pneumonia due to Escherichia coli |

| Code | Description (ICD-10-AM) |
|---|---|
| J156 | Pneumonia due to other Gram-negative bacteria |
| J157 | Pneumonia due to Mycoplasma pneumoniae |
| J158 | Other bacterial pneumonia |
| J159 | Bacterial pneumonia, unspecified |
| J168 | Pneumonia due to other specified infectious organisms |
| J170 | Pneumonia in bacterial diseases classified elsewhere |
| J171 | Pneumonia in viral diseases classified elsewhere |
| J172 | Pneumonia in mycoses |
| J173 | Pneumonia in parasitic diseases |
| J178 | Pneumonia in other diseases classified elsewhere |
| J180 | Bronchopneumonia, unspecified |
| J181 | Lobar pneumonia, unspecified |
| J182 | Hypostatic pneumonia, unspecified |
| J188 | Other pneumonia, organism unspecified |
| J189 | Pneumonia, unspecified |
| J200 | Acute bronchitis due to Mycoplasma pneumoniae |
| J201 | Acute bronchitis due to Haemophilus influenzae |
| J202 | Acute bronchitis due to streptococcus |
| J203 | Acute bronchitis due to coxsackievirus |
| J204 | Acute bronchitis due to parainfluenza virus |
| J205 | Acute bronchitis due to respiratory syncytial virus |
| J206 | Acute bronchitis due to rhinovirus |
| J207 | Acute bronchitis due to echovirus |
| J208 | Acute bronchitis due to other specified organisms |
| J209 | Acute bronchitis, unspecified |
| J210 | Acute bronchiolitis due to respiratory syncytial virus |
| J211 | Acute bronchiolitis due to human metapneumovirus |
| J218 | Acute bronchiolitis due to other specified organisms |
| J219 | Acute bronchiolitis, unspecified |
| J22 | Unspecified acute lower respiratory infection |
| J440 | Chronic obstructive pulmonary disease with acute lower respiratory infection |
| J441 | Chronic obstructive pulmonary disease with acute exacerbation, unspecified |

*ICD-10-AM codes were used to identify the study outcome (International Statistical Classification of Diseases and Related Health Problems, Tenth Revision, Australian Modification). Codes are shown without decimal points.*

##### Table S2 – Model implementations and settings

| Model | Implementation | Explicit settings |
|---|---|---|
| LGB | LightGBM binary classifier | 100 trees; gain importance; is_unbalance=True; one thread; seed 20273112 |
| XGB | XGBoost classifier | 100 trees; scale_pos_weight=n_negative/n_positive; one thread; seed 20273112 |
| Tree (lgb) | LightGBM binary classifier | One boosting iteration; otherwise the specified LGB settings |
| Tree (xgb) | XGBoost classifier | One boosting iteration; otherwise the specified XGB settings |
| LR | scikit-learn logistic regression | Balanced class weights; seed 20273112 |
| RF | YDF random forest | 100 trees; numerical-column discretisation; positive-record weight=n_negative/n_positive and negative-record weight=1 |

Unspecified options use package defaults. StandardScaler is fitted in the training data and then applied to the test data. Ratios for class balancing are calculated from the training sets. Cross-validation groups people by demographic profile, using seed 20273112 to sample and allocate profile representatives.

##### Table S3 – Full variable set model performance

| Model | CV AUROC | CV APS | Next-year AUROC | Next-year APS |
|---|---|---|---|---|
| LGB | 0·8430 (0·0095) | 0·1283 (0·0098) | 0·8429 (0·0098) | 0·1267 (0·0108) |
| XGB | 0·8415 (0·0091) | 0·1285 (0·0101) | 0·8423 (0·0096) | 0·1277 (0·0112) |
| Tree (xgb) | 0·8244 (0·0100) | 0·1054 (0·0088) | 0·8243 (0·0108) | 0·1039 (0·0089) |
| Tree (lgb) | 0·8272 (0·0097) | 0·0811 (0·0088) | 0·8268 (0·0103) | 0·0799 (0·0091) |
| LR | 0·7589 (0·0083) | 0·0748 (0·0060) | 0·7579 (0·0088) | 0·0735 (0·0061) |
| RF | 0·8240 (0·0097) | 0·0720 (0·0083) | 0·8238 (0·0104) | 0·0711 (0·0095) |

Values are means (sample SD across periods). CV gives equal weight to eight annual means, each based on five folds. Next-year evaluation gives equal weight to seven adjacent-year scores. SD describes annual variation, not a confidence interval. The full set of variables comprises demographics, FV, M3, and P3. FV denotes influenza vaccination status; AUROC denotes area under the receiver operating characteristic curve; APS denotes average precision score.

##### Table S4 – Average precision by variable set

| Variables | LGB CV | XGB CV | LGB next year | XGB next year |
|---|---|---|---|---|
| Demo | 0·0720 | 0·0713 | 0·0698 | 0·0696 |
| Demo + FV | 0·0774 | 0·0768 | 0·0750 | 0·0751 |
| Demo + M3 | 0·1034 | 0·1021 | 0·1012 | 0·1011 |
| Demo + P3 | 0·0998 | 0·0996 | 0·0974 | 0·0977 |
| Demo + M3 + P3 | 0·1158 | 0·1154 | 0·1136 | 0·1139 |
| Demo + FV + M3 | 0·1144 | 0·1135 | 0·1123 | 0·1126 |
| Demo + FV + P3 | 0·1117 | 0·1124 | 0·1100 | 0·1108 |
| Demo + FV + M3 + P3 | 0·1283 | 0·1285 | 0·1267 | 0·1277 |

Scores are equal-period means: eight annual CV means or seven next-year evaluations. Demo comprises age, sex, DEP, and five non-exclusive ethnicity indicators. DEP denotes deprivation score; FV denotes influenza vaccination status. M3 and P3 are continuous historical morbidity indices. LGB denotes LightGBM and XGB denotes XGBoost.

**Table S5 – AUROC by variable set**

| Variables | LGB CV | XGB CV | LGB next year | XGB next year |
|---|---|---|---|---|
| Demo | 0·7856 | 0·7830 | 0·7854 | 0·7838 |
| Demo + FV | 0·7891 | 0·7866 | 0·7887 | 0·7872 |
| Demo + M3 | 0·8259 | 0·8233 | 0·8257 | 0·8241 |
| Demo + P3 | 0·8297 | 0·8279 | 0·8296 | 0·8285 |
| Demo + M3 + P3 | 0·8400 | 0·8382 | 0·8398 | 0·8389 |
| Demo + FV + M3 | 0·8283 | 0·8259 | 0·8281 | 0·8266 |
| Demo + FV + P3 | 0·8331 | 0·8315 | 0·8330 | 0·8323 |
| Demo + FV + M3 + P3 | 0·8430 | 0·8415 | 0·8429 | 0·8423 |

Scores are equal-period means: eight annual CV means or seven next-year evaluations. Demo comprises age, sex, DEP, and five non-exclusive ethnicity indicators. DEP denotes deprivation score; FV denotes influenza vaccination status. M3 and P3 are continuous historical morbidity indices. LGB denotes LightGBM and XGB denotes XGBoost.

**Table S6 – Increments associated with vaccination**

| Variables before FV | Metric | LGB CV | XGB CV | LGB next year | XGB next year |
|---|---|---|---|---|---|
| Demo | APS | 0·0054 | 0·0055 | 0·0052 | 0·0055 |
| Demo + M3 | APS | 0·0110 | 0·0114 | 0·0111 | 0·0115 |
| Demo + P3 | APS | 0·0119 | 0·0128 | 0·0127 | 0·0131 |
| Demo + M3 + P3 | APS | 0·0124 | 0·0131 | 0·0130 | 0·0138 |
| Demo | AUROC | 0·0035 | 0·0036 | 0·0033 | 0·0034 |
| Demo + M3 | AUROC | 0·0024 | 0·0026 | 0·0024 | 0·0025 |
| Demo + P3 | AUROC | 0·0034 | 0·0036 | 0·0035 | 0·0037 |
| Demo + M3 + P3 | AUROC | 0·0030 | 0·0033 | 0·0030 | 0·0034 |

Each value is the mean score with FV minus the score without FV. Comparisons are matched by year and fold for CV, and by training/evaluation year pair for temporal evaluation. Full-precision scores are differenced before rounding. FV denotes influenza vaccination status; Demo and the other abbreviations follow tables S3 and S4. These are model-performance differences, not estimates of vaccine effectiveness.

**Table S7 – Mean gain and split importance**

| Feature | LGB gain | LGB split | XGB gain | XGB split |
|---|---|---|---|---|
| Age | 44·69 | 35·43 | 42·40 | 30·21 |
| M3 | 36·58 | 13·61 | 32·51 | 12·50 |
| P3 | 14·91 | 19·77 | 15·53 | 17·60 |
| DEP | 1·45 | 15·98 | 3·74 | 24·15 |
| FV | 1·12 | 3·14 | 1·93 | 2·96 |
| Sex | 0·17 | 3·88 | 0·72 | 3·25 |
| Asian | 0·20 | 1·82 | 0·57 | 2·15 |
| Europe | 0·20 | 1·98 | 0·72 | 2·18 |
| Māori | 0·31 | 2·08 | 0·86 | 2·06 |
| Pacific | 0·36 | 1·69 | 0·89 | 1·68 |
| MELAA | 0·02 | 0·62 | 0·12 | 1·26 |

Values are percentages averaged over five folds and eight years. Importance is normalised separately within each model and measure. Gain denotes total split gain and split denotes split frequency. DEP denotes deprivation score; FV denotes influenza vaccination status. Europe and Pacific denote European and Pacific peoples ethnic groups, respectively. MELAA denotes Middle Eastern, Latin American, and African. Ethnicity indicators are non-exclusive.

### Supplementary figure S1 SHAP profiles for the additional application year

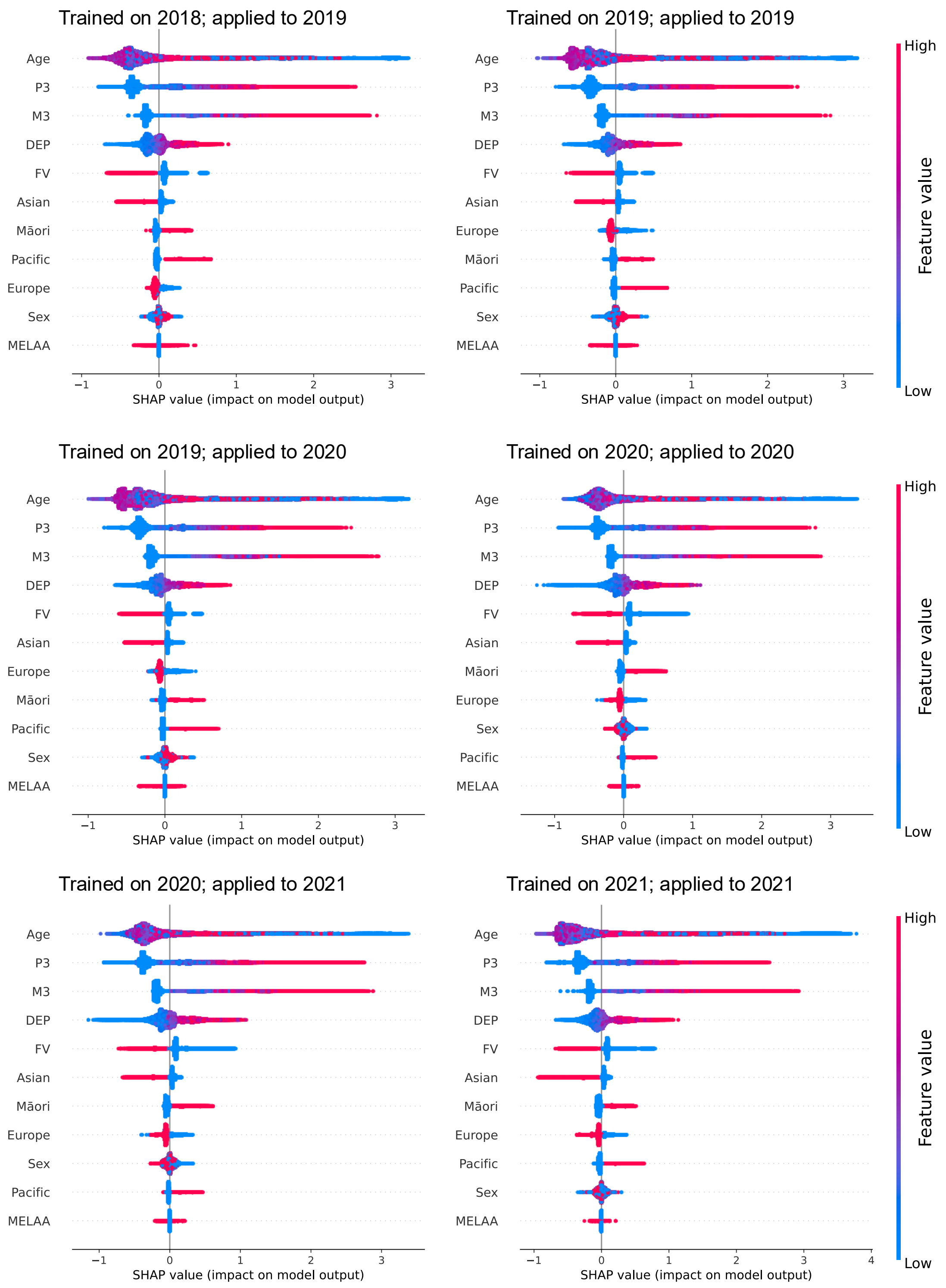

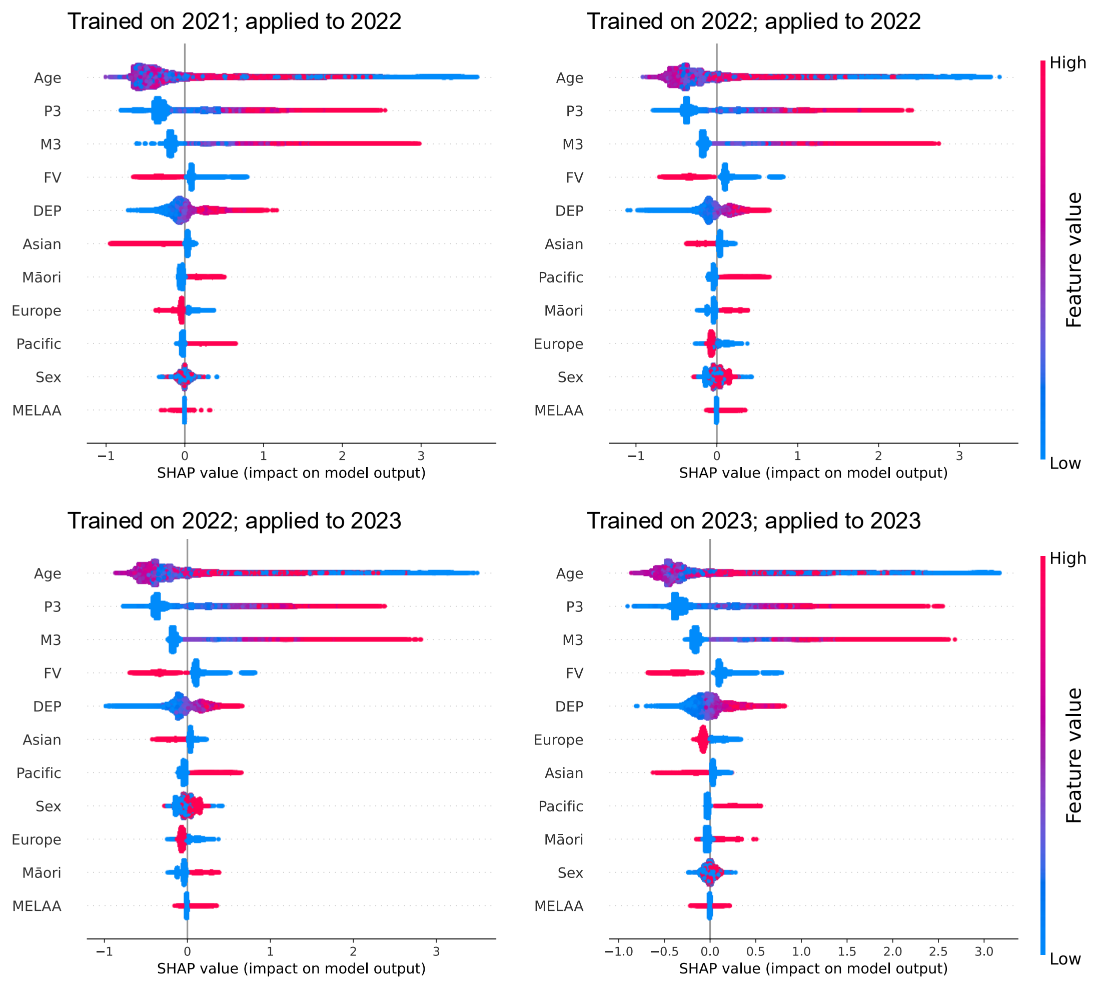


SHapley Additive exPlanations (SHAP) beeswarm profiles for full-variable LightGBM models in application years not displayed in **Figure 5**. Subpanel titles will identify the training and application years. Same-year profiles describe records from the model-fitting dataset; next-year profiles apply the preceding year's fitted model. Positive and negative SHAP values indicate higher and lower model output relative to the reference expectation. Colour indicates low (blue) to high (red) variable values. SHAP values retain their model-output scale. DEP denotes deprivation score; FV, influenza vaccination status; Europe and Pacific, European and Pacific peoples, respectively; MELAA, Middle Eastern, Latin American, and African. Ethnicity indicators are non-exclusive.
